# A serological assay to detect SARS-CoV-2 antibodies in at-home collected finger-prick dried blood spots

**DOI:** 10.1101/2020.05.29.20116004

**Authors:** Donna Grace Karp, Kenneth Danh, David Seftel, Peter V. Robinson, Cheng-ting Tsai

## Abstract

Accurate surveillance of coronavirus disease 2019 (COVID-19) incidence requires large-scale testing of the population. Current testing methods require in-person collection of biospecimens by a healthcare worker, limiting access of individuals who do not have access to testing facilities while placing both patients and healthcare workers at risk of exposure to infection. We report the development and validation of a at-home finger-prick dried blood spot collection kit and an analysis method. We demonstrated 100% sensitivity and specificity using at-home collected specimens across the US. Such methods may facilitate the conduct of unbiased serosurveys within hard to reach populations and help reduce the sample collection burden of serological testing on both health care systems and individuals alike.

## INTRODUCTION

Severe acute respiratory syndrome coronavirus 2 (SARS-CoV-2), formerly known as 2019-nCoV, is an enveloped, single-stranded RNA virus in the family Coronaviridae, genus Betacoronavirus^1–3^. At the time of writing, SARS-CoV-2 pandemic has led to more than 4.3 million confirmed cases globally and claimed approximately 300,000 lives.

While direct detection of the virus by RNA or antigen assays remains the standard for confirming acute SARS-CoV-2 infection, serological antibody testing is of increasing importance for many reasons^4–8^. Antibody tests can complement RNA tests by improving the diagnostic yield among suspected cases that have false negative RNA/antigen tests due to sampling or assay technical limitations. Furthermore, serological assays are also important for assessing asymptomatic infection in close contacts, identifying individuals with resolved infection as potential contributors of convalescent plasma therapy, enabling estimation of population infection prevalence and illuminating our understanding of contributing factors conferring immunity to reinfection.

All currently approved serological tests require blood specimens collected inperson by a healthcare worker either through phlebotomy or finger-prick. The need for in-person collection limits access to testing for individuals who may not be able to get to a testing center due to distance, disability or inconvenience. The in-person collection process is labor intensive, driving up testing costs and diverting healthcare resources from the care of ill individuals. In-person collection processes carry an increased risk of infection due to potential exposure to a confluence of contagious individuals and consumes large quantities of valuable personal protective equipment (PPE).

In principle, rapid tests such as lateral flow assays may at some point be approved for at-home testing. However, lateral flow tests have shown an alarmingly wide performance dispersion even when performed by trained personnel in controlled clinical settings, with many tests exhibiting inadequate sensitivity and specificity^9^. The accuracy of lateral flow tests for at-home testing will require significant improvement and extensive additional validation before they can be trusted as clinically and epidemiologically useful. Integrity in self-reporting of the results of home-administered lateral flow tests also poses major challenges.

Here, we report the development of an at-home finger-prick dried blood spot collection kit and an accompanying laboratory test to elute and accurately detect antibodies in this convenient sample type with 100% concordance to their clinical status. For this study, we created standardized dried blood spot collection kits for reciprocal mailing to and from the homes of potential participants. These kits contained lancets for self-collection of blood and dried blood spot cards to capture and preserve the specimens for transport. The kits were then mailed back to a central laboratory for extraction and analysis. This type of home-to-lab sourced samples can provide the high quality and consistency of a reference lab based serological test without exposing patients or healthcare workers to unnecessary risks.

## METHODS

### Materials

The SARS-CoV-2 spike protein (S1) containing amino acids 1–674 with an Fc-tag at the C-terminus (#31806), expressed in HEK293 cells, was purchased from the Native Antigen Company (Oxford, United Kingdom). Oligonucleotides used in the study were custom ordered from Integrated DNA Technologies (Coralville, IA). Platinum Taq polymerase (#10966026), SYBR qPCR 2X master mix (#4385610), Dithiothreitol (DTT #202090) and sulfo-SMCC (#22122) were purchased from Thermo Fisher (Waltham, MA). DNA ligase (#A8101) was purchased from LGC (Teddington, United Kingdom). Other reagents are detailed in the method sections as appropriate.

### Human specimens used in the study

Individuals who tested positive for SARS-CoV-2 RNA by RT-PCR between March – April 2020 were recruited to the study via social media. Healthy donors without SARS-CoV-2 symptoms were enrolled as negative controls. All participants provided informed consent under the Western IRB approved IRB protocol #20180015 to Enable Biosciences. Dried blood spot collection kits were mailed to each participant for at-home sample collection. We requested two to five dried blood spots from each participant. Completed dried blood spots were dropped off in standard United States Postal Service (USPS) blue drop boxes for ground return shipment to Enable Biosciences. The entire process was done at ambient temperature without a cold chain.

A total of 56 self-collected finger-prick dried blood spot specimens were received. Negative control specimens were collected from 25 donors, and positive specimens were collected from 31 COVID-19 patients. The patients were deemed infected if symptomatic and tested positive by RT-PCR (N = 30) or by validated serum antibody tests (N = 1).

In addition to dried blood spots, K2- EDTA plasma specimens were collected by phlebotomy from 8 selected participants (4 from COVID-19 patients and 4 from health controls).

### Dried blood spot collection kit

The dried blood spot collection kit contained a sample collection instruction sheet, disposable lancets, a dried blood spot card, alcohol pads, Band Aids and a pre-printed return label. The participant was asked to clean the collection site with the sterile alcohol pad and then use the disposable lancet to prick the finger. Finger-prick blood was then carefully dropped onto each dried blood spot card until a complete circle was filled. A minimum of two spots per participant was requested. The kit contained multiple lancets in case a single prick proved insufficient to generate multiple spots. Given that the SARS-CoV-2 virus is rarely detected in the blood of recovered donors or donors with mild to moderate symptoms, the dried blood spot specimens presented a minimal biohazard risk^10,11^. Furthermore, dried blood spots were fully dried prior to shipment, thus further lowering the risk profile. The dried blood spot return kits were transported by standard postal mail service with UN3373 Biological Substance Category B Labels affixed. Once received, the dried blood spot cards were stored at 2–8°C until analyzed.

### Detection of SARS-CoV-2 antibodies in dried blood spots

The dried blood spot analysis comprised elution and testing steps. For elution, six 3mm discs were punched from a single dried blood spot. In rare circumstances where a dried blood spot was not completely filled, six 3mm discs were punched from multiple spots. Then, the discs were incubated with 1000 μL of elution buffer for 90 min on a heat shaker at 37°C for elution. Then, the eluent was concentrated with a 100 kDa molecular weight cut-off column (MWCO) for 8min at 14,000 rcf.

For the testing, the eluent was assayed using a modified Antibody Detection by Agglutination-PCR (ADAP) protocol^12–14^. Briefly, 8 μL of eluent was incubated with 8 μL of 1 femtomole of SARS-CoV-2 spike protein S1 subunit-DNA conjugate pair mixtures at 37 °C for 30 min. If present, the SARS-CoV-2 antibodies in the specimen engage with S1-DNA conjugate to agglutinate into a dense immune complex. To quantify the degree of agglutination, 116 μL of ligation solution (20 mM Tris, 50 mM KCl, 20 mM MgCl2, 20 mM DTT, 25 μM NAD, 0.025 U/μl ligase, 100 nM connector) was mixed with 4 μL eluent/conjugate mixture and incubated at 30 °C for 15 min. Then, 25 μL of the ligated solution was mixed with PCR master mix that contained primer pairs and polymerase for amplification under standard thermocycling conditions (95 °C for 10 min, 95 °C for 15 sec, 56 °C for 30 sec, 13 cycles). The pre-amplified products were then quantified in a 96-well qPCR plate. SYBR green-based qPCR was performed on a Bio-Rad CFX96 real-time PCR detection system (95 °C for 10 min, 95 °C for 30 sec, 56 °C for 1 min, 40 cycles). As a result of agglutination, specimens harboring high quantities of antibodies will have large quantities of amplifiable DNA, thus exhibiting a strong qPCR signal (low Ct). In contrast, samples without SARS-CoV-2 antibodies will have few DNA amplicons, thus yielding a weaker qPCR signal (high Ct). Instead of using the common cycle threshold (Ct) as readout, the ADAP assay readout ΔCt is defined as the Ct value of a blank control minus the Ct of the actual specimen. The magnitude of the ΔCt is proportional to the amplicon concentration in the PCR plate well, which in turn is proportional to the amount of antibody present in the sample. The ΔCt offers significant reproducibility since the subtraction of the blank control Ct and the sample Ct cancels out any potential drift across runs.

To ensure the quality of the process, one positive and one negative control dried blood spots cards were always analyzed concurrently. The control cards went through the entire elution and testing process. The analysis was only valid if the positive control cards tested positive and the negative cards tested negative.

## RESULTS

### Cohort characteristics

We used social media (e.g. Facebook, Twitter, Medium) and community referrals to recruit volunteers for the study. We aimed to enroll both COVID-19 patients (positive controls) and healthy donors (negative controls) to validate the sensitivity and specificity of the serological test using self-collected dried blood spots. The COVID-19 patients were eligible to participate in the study if they presented with COVID-19 symptoms and were confirmed positive by either RT-PCR for SARS-CoV-2 RNA and/or a validated serum/plasma serological tests for SARS-CoV-2 antibodies. The control patients were included in the study if they had no symptoms and/or no known exposure to SARS-CoV-2 and/or tested negative by validated serum/plasma serological tests for SARS-CoV-2 antibodies.

Eligible participants received a dried blood spot mailer kit that included comprehensive instructions for self-collection of finger-prick spots. The mailer kit contained a pre-printed return label to ship the kit back to the Enable Biosciences laboratory in South San Francisco by USPS.

A total of 56 specimens were received for the study, where 31 were from COVID-19 patients and 25 were from healthy donors. The patient demographics were summarized in **Table 1**. Notably, several donors were from out-of-state locations distant from the Enable Biosciences facilities. Since the dried blood spots were returned via ground shipping at ambient temperature, the inclusion of such donors allowed us to investigate the potential impact of long-distance transportation on sample quality and analytical performance.

**Table 1.** Study participant demographics. For 7 control patients, the age information is not provided.

|  |  | COVID-19 | Healthy control |
| --- | --- | --- | --- |
| Number of donors |  | 31 | 25 |
| Gender |  | 20 Female | 16 Female |
| Age (yr) |  | 24-86 | 15-59* |
| Distance to Enable Biosciences Lab (miles) | <100 | 20 | 21 |
|  | 100-1500 | 0 | 2 |
|  | >1500 | 11 | 2 |
| Laboratory confirmation | PCR | 30 | 0 |
|  | Serum/Plasma antibody test | 1 | 4 |

### Detection of SARS-CoV-2 antibody in self-collected finger-prick dried blood spots

We first evaluated the clinical sensitivity and specificity of self-collected finger-prick dried blood spots. Among 31 COVID-19 patients, 31 tested positive for SARSCoV-2 antibodies. For 25 healthy donors, 25 tested negative for SARS-CoV-2 antibodies. The preliminary results indicated 100% sensitivity and 100% specificity of mailer-based finger-prick dried blood spot specimens (**Fig. 1**). The study is still ongoing to expand the cohort size.

**Figure 1.**
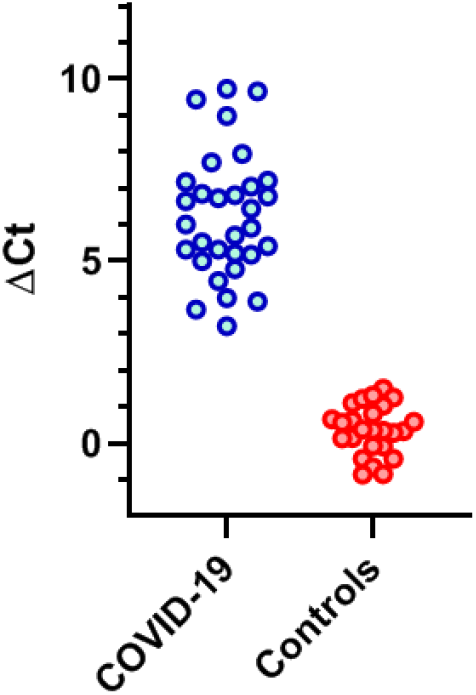
SARS-CoV-2 antibodies levels in self-collected finger-prick dried blood spots. Dried blood spot eluents were tested by the ADAP method for antibodies against the S1 protein of SARS-CoV-2. Signals were coded blue for COVID-19 and red for control donors. The y-axis is the signal output from ADAP, calculated by subtracting the Ct value of the specimen to the blank control (buffer C).

In addition to clinical performance, we also evaluated the signal distribution between samples collected in immediate proximity to the testing laboratory and those from distant areas. For COVID-19 patients, 20 specimens were collected within 100 miles of testing labs, while 11 specimens were collected from locations over 1500 miles away. The signal distribution between these two groups did not reach statistical significance (**Fig. 2**). Similar observations were made for the healthy control patient groups. The actual transit time of the completed kits, irrespective of the individual’s physical distance away from the lab, ranged from 0–5 days. 88% of all test kits arrived within 3 days of the person mailing the completed kit back to Enable Biosciences using either USPS Priority or First-Class Mail. The data demonstrated excellent dried blood spot specimen integrity even after long-range transportation at ambient temperatures.

**Figure 2.**
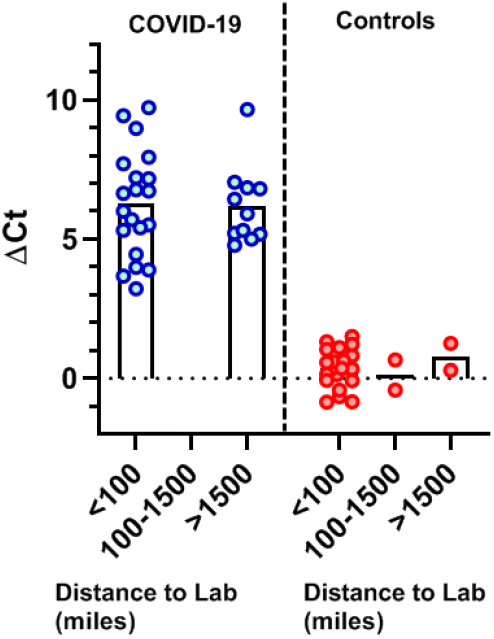
Finger-prick dried blood spot signals by distance to the testing lab. The self-collected finger-prick samples were grouped into short distance (<100 miles), moderate distance (100–1500 miles) and long distance (>1500 miles) based on the geographical distance between the donor’s resident city and South San Francisco (the site of the testing lab).

### Correlation of SARS-CoV-2 antibody signals between sample types

In addition to sensitivity and specificity, it is critical to compare the antibody signals in self-collected finger-prick dried blood spots to venipuncture-based blood samples. The lancets used in self-collected finger-prick samples are shorter than the needles used for venipuncture. Untrained individuals collecting at home may use a wide range of lancing techniques and differing puncture sites. As a result, finger-prick blood could conceivably be partially diluted by extra-venous tissue fluids.

To evaluate the impact of these possibilities, we collected K2-EDTA plasma samples by standard phlebotomy from 4 COVID-19 patients and 4 healthy controls. The plasma samples were tested using the standard ADAP assay for SARS-CoV-2 antibodies. The data showed that the plasma signals were highly consistent and well correlated with those from self-collected sample types (**Fig. 3**), effectively eliminating the theoretical sampling concerns of at-home collected dried blood spot samples for SARS-CoV-2 antibody measurement.

**Figure 3.**
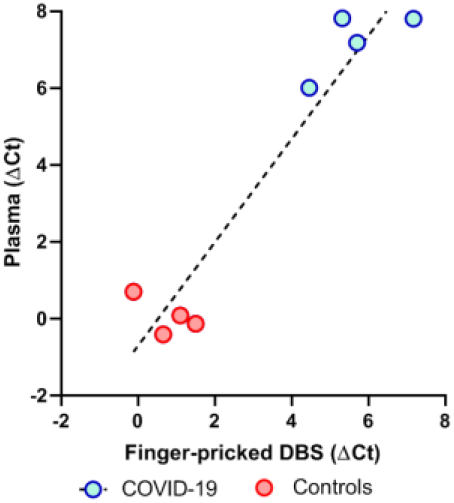
Correlation of signals between finger-prick dried blood spots and venipuncture plasma samples. (R = 0.96).

### DISCUSSION

Serological testing plays a significant role in the management of SARS-CoV-2 infection. First, RT-PCR may produce false negative results in the setting of low viral loads. Antibodies could serve as a complementary marker to boost the accuracy of RTPCR testing in the waxing and waning phases of viral infection. Secondly, the presence of antibodies indicates past exposure to SARS-CoV-2. Thus, antibody-based serosurveys are key to measuring disease prevalence at the population level. This data is critical for accurate characterization of essential parameters such as fatality rates, basic reproduction number (Ro) and others^15^. Finally, convalescent plasma has been shown as a safe and promising therapy to mitigate disease^16,17^. Serological testing can help identify eligible donors for further antibody characterization by methods such as neutralization assays.

However, the constraint of phlebotomy for sample collection for current serological tests has curtailed these potential benefits. Many at-home collection RTPCR RNA tests have received regulatory approvals. To yield a complete picture of SARS-CoV-2 infection, it is highly desirable to pair suitable molecular diagnostic with self-collected serological tests

The dried blood spot collection kit and testing method reported in this study have the potential for wide-spread use. Obviating phlebotomy could improve both testing safety and testing access, while reducing the sample collection burden on severely stressed healthcare systems. The stability of dried blood spots at ambient temperature for extended time periods permits standard mail parcel transportation service use without requiring cold chain preservation measures. Importantly, mailer-based self-collected samples may reduce bias and improve participation within underserved or sequestered communities by reaching populations beyond those accessed by serosurveys that rely solely on collection at drive-through clinics.

Since the dried blood spot modality has the potential for wide scale screening, it is critical to ensure the specificity of this test. The use of ADAP methods for dried blood spot eluent testing may have an additional unique advantage in that the ADAP method consumes a very small volume of eluent (e.g. 8 μL) per test. It is thus possible to conserve samples to support further analyses via a comprehensive reflex testing algorithm to further enhance test accuracy. For instance, it is potentially feasible to conduct the primary screening of the eluent by detecting only anti-S1 protein antibodies. Samples that test positive can then be reflexed for confirmatory testing for anti-N protein antibody without having to elute additional samples from another spot.

In summary, we report a self-collection kit for at-home finger-prick dried blood spot collection with a method to elute and test the specimen that show comparable analytical performance to venipuncture-derived blood samples. If proven successful at large scale, this method can greatly facilitate the conduct of unbiased serosurveys within hard to reach populations and help reduce the sample collection burden of serological testing on both health care systems and individuals alike.

## Data Availability

All reagents and data from this study will be made available upon reasonable request from the corresponding author.

## ACKNOWLEDGEMENTS

We thank all patients, donors and health care staff who participated in this study. We thank Corleone Delaveris and Melissa Gray for assistance on execution of the study. The study is funded in part by NIH SBIR 2R44DK110005–02 and 2RAI141118–02 to Enable Biosciences. The content is solely the responsibility of the authors and does not necessarily represent the official views of the NIH.

